# Post-discharge mortality in suspected pediatric sepsis: insights from rural and urban healthcare settings in Rwanda

**DOI:** 10.1101/2024.11.08.24316988

**Authors:** Christian Umuhoza, Anneka Hooft, Cherri Zhang, Jessica Trawin, Cynthia Mfuranziza, Emmanuel Uwiragiye, Vuong Nguyen, Aaron Kornblith, Nathan Kenya Mugisha, J Mark Ansermino, Matthew O. Wiens

## Abstract

Post-discharge death is increasingly recognized as an important contributor to pediatric mortality in sub-Saharan Africa. To address morbidity and mortality during this period, a representative evidence base is needed to inform resource prioritization, policy, and guideline development. To date, no studies have been conducted in Rwanda, limiting understanding of post-discharge mortality in this region. We conducted a prospective cohort study of children ages 0-60 months in two Rwandan hospitals, one rural (Ruhengeri) and one urban (Kigali), from May 2022 to February 2023. We collected clinical, laboratory, and sociodemographic data on admission and follow-up data on vital statistics at 2-, 4-, and 6-months post-discharge. Of 1218 children enrolled, 115 (9.4%) died, with half occurring in-hospital (n=57, 4.7%) and half after discharge (n=58, 4.7%). Post-discharge mortality was lower in the 6-60-month cohort (n=30, 3.5%) than in the 0-6-month cohort (10%) and higher in Kigali (n=37, 10.3%) vs. Ruhengeri (n=21, 2.7%). Median time to post-discharge death was 38 days (IQR: 16-97.5) in the 0–6-month cohort and 33 days (IQR: 12-76) in the 6– 60-month cohort. In the 0-6 months cohort, malnutrition (weight-for-age z-score <-3) was associated with increased odds of post-discharge death (aOR 3.31, 95% CI 1.28-8.04), while higher maternal education was protective (aOR 0.15, 95% CI 0.03-0.85). Significant factors associated with post-discharge death in the 6-60-month cohort included an abnormal Blantyre Coma Scale (aOR 3.28, 95% CI 1.47-7.34), travel time to care >1 hour (aOR 3.54, 95% CI 1.26-9.93), and referral for higher levels of care (aOR 4.13, 95% CI 1.05-16.27). Children aged <2 months exhibited the highest cumulative mortality risk. Post-discharge mortality among Rwandan children remains a significant burden, necessitating targeted interventions for post-discharge care and follow up to reduce mortality.

## Introduction

Pediatric mortality following hospital discharge is often overlooked aspect of child health in Sub-Saharan Africa (SSA). Despite significant decreases in in-hospital mortality rates in pediatric patients in SSA, the early post-discharge period is an especially vulnerable time marked by an increased risk of death, primarily within the first six months [3–5]. An in-depth understanding of the many complex factors contributing to pediatric post-discharge mortality is a critical first step in the design and implementation of effective targeted interventions to reduce the burden of sepsis and improve outcomes in low-resource settings.

Existing research from low- and middle-income country (LMIC) settings has provided valuable insights into the epidemiology and risk factors associated with post-discharge mortality in children. The multi-county Child Health and Mortality Prevention Surveillance Network (CHAIN) study, evaluated the causal structure of post-discharge mortality, outlining its complex nature, as it pertains to social, nutritional, and illness-related vulnerability, and identified the importance of malnutrition in post-discharge mortality [6]. In Uganda, the Smart Discharges studies have emphasized that clinical, socioeconomic, behavioral, and lab-based risk factors present on admission can be used for risk stratification of children to inform a more personalized approach to post-discharge care [5,7]. These risk factors have been used to create predictive algorithms to identify children at highest risk of post-discharge death, in whom low-cost interventions based on this individual risk can be applied to reduce mortality [5,8]

Like most of sub-Saharan Africa, Rwanda has seen dramatic reductions in child mortality, likely a cumulative result of several different, national-level interventional programs [9]. These include interventions such as introduction of community health insurance, performance-based pay for providers [10], geographical accessibility improvements [11], health system strengthening partnerships [12], nurse mentorship programs [13], community health workers programs [14], and data-driven quality improvement initiatives [15]. Despite a growing body of evidence describing the risks and burden of post-discharge death in children treated for infections [3], its epidemiology in Rwanda has not yet been evaluated.

With the complex interplay of health systems, population-, and individual-level risk factors, Rwanda not only provides a unique setting for investigating post-discharge mortality among children [14]. Given the existing healthcare infrastructure improvements in Rwanda, a better understanding of disparities in post-discharge mortality may further enhance their efficacy. This study aimed to investigate the epidemiology of post-discharge mortality in children admitted with suspected sepsis in Rwanda and to identify the key risk factors.

## Materials and Methods

### Study Design and Setting

This prospective cohort study was conducted at two Rwandan hospitals. Situated in the Northern Province, Ruhengeri Referral Hospital operates as the main referral center and the only district hospital in Musanze District, serving a largely rural population with a catchment area nearing 500,000. The hospital has four total ICU beds and no pediatric-specific ICU capacity. The second, the University Teaching Hospital of Kigali (CHUK), located in Nyarugenge District, Kigali City, is the largest hospital in the country and serves as Rwanda’s primary referral center, with a capacity of 483 beds. It also serves as a teaching hospital and center for clinical research for multiple medical schools and provides technical assistance to the surrounding district hospitals.

### Participant Recruitment and Selection Criteria

We enrolled a cohort of children ages 0-60 months between May 2022 and February 2023. These groups were stratified into 0-6 months and 0-60 month sub-cohorts given prior variability in risk predictors and model development specific to these age groups informed by prior studies [16,17]. Inclusion criteria included: any child within this age group admitted with suspected sepsis, defined as suspected or proven infection by treating clinical team. We excluded children living outside the hospital service area, admitted for short-term observation (less than 24 h), or treated for trauma or non-infectious illness. Previous research in similar settings in Uganda demonstrated that 90% of children admitted with a confirmed or suspected infection met the International Pediatric Sepsis Consensus Conference criteria for sepsis [5,18], Written informed consent was obtained from the parents or legal guardians of all participants. Children whose parents or caregivers refused to participate were excluded from the study.

### Data Collection Procedures

Data were collected at admission, discharge, and at 2, 4, and 6-months post-discharge. A research nurse collected information on clinical history and evaluation, laboratory findings, and sociodemographic characteristics, which mirrored the methodologies used in a similar study conducted in Uganda, ensuring consistency and comparability across studies [5]. All data collection instruments are accessible via the Smart Discharges study dataverse [1]. Data were gathered directly at the point of care using encrypted study tablets and subsequently uploaded to a Research Electronic Data Capture (REDCap) system [19]. We used a combination of telephone interviews and home visits by research field officers for follow-up visits. These follow-ups focused on vital status, health-seeking behaviors, and any readmissions.

### Variables and Measurements

Clinical information collected included vital signs, anthropometric measurements to determine malnutrition status, basic laboratory tests (such as glucose levels, malaria and HIV rapid diagnostic tests [RDTs], hematocrit, and lactate), observed clinical signs and symptoms, comorbidities, and healthcare history, including any prior hospital admissions. We evaluated nutritional status using weight-for-age z-scores based on the World Health Organization (WHO) growth standards [20].

Sociodemographic data included maternal and household details, such as mother’s age, education level, HIV status, household size, use of bed nets, proximity to the health facility, and availability of clean drinking water. Information on the child’s sex was obtained from medical records. At the time of discharge, the study nurses recorded the discharge status (categorized as routine discharge, referral for higher-level care, or unplanned discharge) and feeding status, which were subjectively assessed as feeding well or feeding poorly. Discharge diagnoses were also retrieved from medical records. Field officers contacted caregivers by telephone at 2-, 4-, and 6-months post-discharge to assess the child’s vital status, any instances of seeking medical care after leaving the hospital and details of any readmissions. In cases where contact was lost, we conducted in-person visits to gather this information. In instances where a child passed away following hospital discharge, we performed verbal autopsies to determine the likely cause of death.

### Statistical Analysis

We performed descriptive statistics to characterize baseline clinical, social/maternal, and discharge variables, stratified by age cohort using medians with interquartile ranges for continuous variables and counts with percentages for categorical variables. Multivariate logistic regression models were used to determine risk factors for post-discharge mortality by estimating the odds ratios adjusted for age, sex, and site of enrollment. Post-discharge mortality was treated as a binary outcome, and the site of enrollment was included as a fixed effect because only two sites were included. We also examined the secondary outcome, readmission post-discharge, using descriptive statistics. We estimated the cumulative hazard for mortality and readmission after discharge with Kaplan-Meier survival curves at four predefined age strata (0–<2 months, 2–6 months, >6–24 months, and >24–60 months). We had minimal missing data and addressed these using k-nearest neighbor imputation to ensure the robustness and validity of the results. We conducted all statistical analyses using Stata/MP version 15.0 (StataCorp, College Station, TX, USA), R version 4.1.3, and RStudio version 2022.2.3 (RStudio, Boston, MA, USA).

### Ethical Considerations

The study was approved by several institutional review boards: The University of California, San Francisco (UCSF) on October 8, 2021 (No. 21-34663); the University of British Columbia (UBC) on January 28, 2022 (No. H21-02795), University of Rwanda on December 30, 2021 (No. 411), and University Teaching Hospital of Kigali on January 14, 2022 (No. 005). Informed consent was obtained from the parents or legal guardians of all participants in Kinyarwanda. This manuscript adheres to the STrengthening the Reporting of OBservational studies in Epidemiology (STROBE) statement for cohort studies [21].

## Results

We enrolled 1,218 children over the 9-month study period, of whom 1,161 survived to hospital discharge and 1,127 completed follow up at 6 months’ post-discharge. There were 115 (9.4%) deaths, evenly split between in-hospital (n=57, 4.7%) and the post-discharge period (n=58, 4.7%) (**Figure 1**). Median age of participants was 13.5 months (IQR 6.1-24.7) with 60% of the cohort being male (n=676) (**Table S1**).

**Figure 1:**
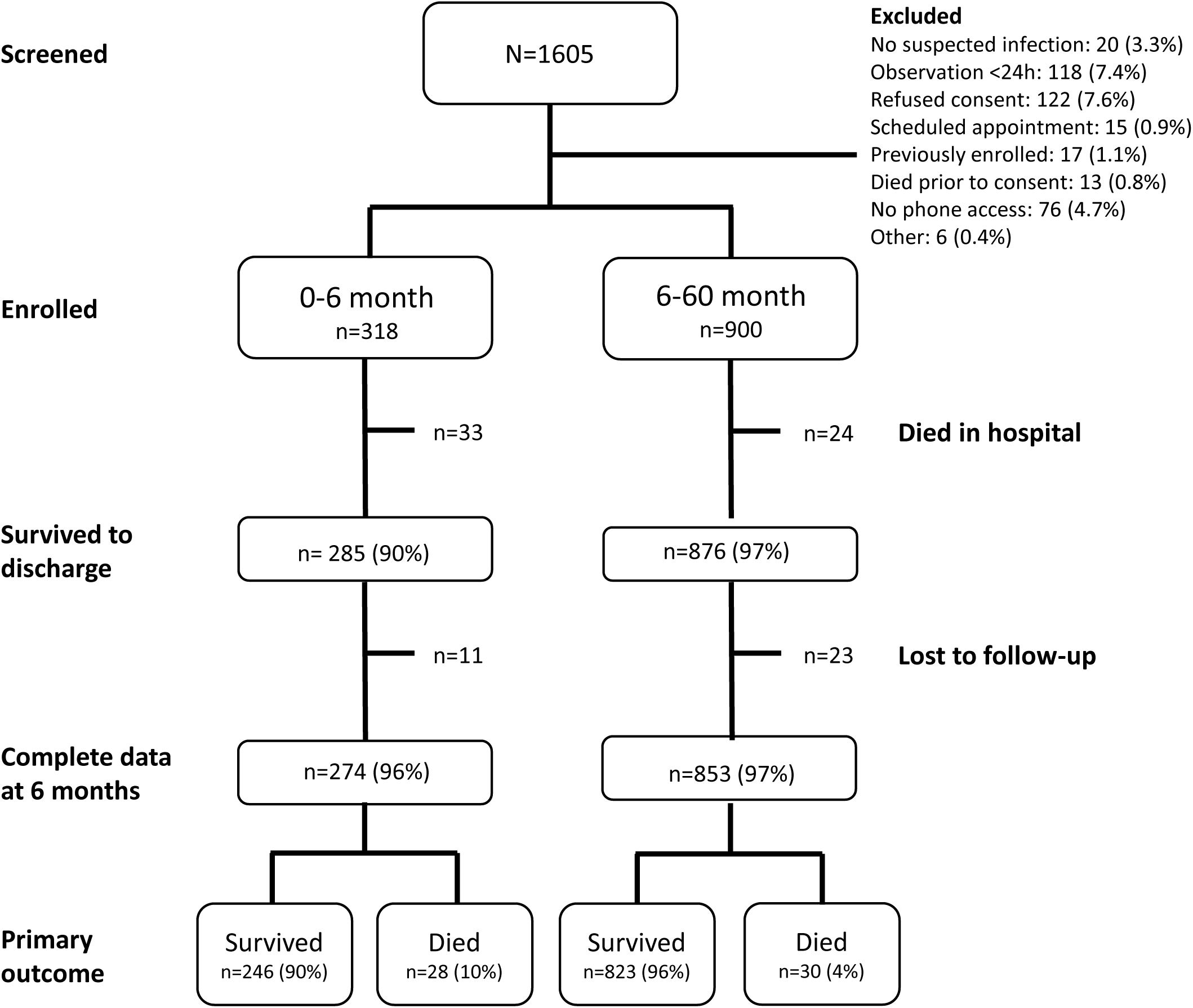
Study flowchart stratified by age group.

Severe malnutrition was common, with 9.1% (n=103) having a weight-for-age z-score (waz) below −3, although this differed significantly between the 0-6 month and 6-60 month age groups **(Table S1, Table 1**). Fewer than half of the children (44.4%, n = 500) had a measured fever on presentation (temperature > 37.5 °C), while 18% (n=208) had measured hypothermia (temperature <36.5 °C). An abnormal Blantyre Coma Scale (BCS) score indicating impaired consciousness was observed in 17.2% of patients (n=194). Only 1.5% (n=17) were malaria positive and and 0.3% (n=3) tested positive for HIV. Anemia, defined as Hemoglobin level <11 g/dL was present in 36.8% (n=415) (**Table S1**). By disposition, 97.3% of all admitted children were routinely discharged, 1.9% and 0.8% referred to higher care and left against medical advice, respectively (**Table S1**). Characteristics of all enrolled children are detailed in **Table S1**, and the characteristics of the two primary age categories are described in **Table 1**.

**Table 1:** Cohort characteristics and disposition, stratified by ages 0-6 months and ages 6-60 months.

|  | 0m to 6m (n=274) | 6m to 60m (n=853) |
| --- | --- | --- |

| Variable | N (%) / Median (IQR) | aOR (95% CI) | N (%) / Median (IQR) | aOR (95% CI) |
| --- | --- | --- | --- | --- |
| **Demographics** |  |  |  |  |
| **Site, n (%)** |  |  |  |  |
| Kigali | 110 (40.2) | reference | 251 (29.4) | reference |
| Ruhengeri | 164 (59.9) | 0.33 (0.15-0.75) | 602 (70.6) | 0.21 (0.10-0.46) |
| **Sex, n (%)** |  |  |  |  |
| Female | 112 (40.9) | reference | 339 (39.7) | reference |
| Male | 162 (59.1) | 0.73 (0.33-1.63) | 514 (60.3) | 0.87 (0.41-1.86) |
| **Age, months\*** | 1.4 (0.6-3.6) | 0.91 (0.72-1.14) | 17.9 (11.3-29.8) | 0.98 (0.95-1.01) |
| **Admission anthropometry** |  |  |  |  |
| **MUAC (mm)\*<sup>1</sup>** | 120 (104-140) | 0.98 (0.96-1.00) | 150 (140-160) | 0.98 (0.97-1.00) |
| <110/<115 | 83 (30.3) | 2.27 (0.65-7.88) | 26 (3.1) | 2.69 (0.70-10.31) |
| 110-120/115-125 | 70 (25.6) | 3.05 (1.14-8.16) | 42 (4.9) | 2.48 (0.68-9.05) |
| >120/ >125 | 121 (44.2) | reference | 785 (92.0) | reference |
| **Weight for age z-score** | -0.9 (-2.2-0.02) | 0.75 (0.62-0.90) | -0.6 (-1.6-0.3) | 0.61 (0.50-0.74) |
| <-3 | 47 (17.2) | 3.21 (1.28-8.04) | 56 (6.6) | 6.52 (2.63-16.16) |
| -3 to -2 | 24 (8.8) | 2.22 (0.64-7.66) | 92 (10.8) | 3.17 (1.17-8.61) |
| >-2 | 203 (74.1) | reference | 705 (82.7) | reference |
| **Admission clinical assessment** |  |  |  |  |
| **SpO2, %\*** | 95 (88-98) | 0.97 (0.92-1.02) | 94 (88-97) | 0.99 (0.95-1.04) |
| **Heart rate** | 150 (137-162) | 1.02 (1.00-1.04) | 140 (125-154) | 0.99 (0.98-1.01) |
| **Respiratory rate\*** | 46 (40-55) | 1.01 (0.98-1.04) | 40 (34-46) | 1.01 (0.98-1.05) |
| **Temperature\*** | 36.9 (36.5-37.9) |  | 37.4 (36.7-38.2) |  |
| < 36.5 | 63 (23.0) | 1.01 (0.38-2.71) | 145 (17.0) | 0.40 (0.09-1.82) |
| 36.5-37.5 | 123 (44.9) | reference | 296 (34.7) | Reference |
| >37.5 | 88 (32.1) | 0.53 (0.20-1.43) | 412 (48.3) | 0.83 (0.38-1.80) |
| **Abnormal BCS\*** | 74 (27.0) | 1.05 (0.44-2.54) | 120 (14.1) | 3.28 (1.47-7.34) |
| **HIV positive\*** | 1 (0.4) | - | 2 (0.2) | 11.42 (0.67-194.83) |
| **Positive malaria test** | 3 (1.1) | - | 14 (1.6) | - |
| **Hemoglobin, g/dl\*** | 12 (10.7-13.5) | 0.83 (0.71-0.97) | 11.3 (10.2-12) | 0.84 (0.71-1.00) |
| No anemia:<br>≥11g/dL | 192 (70.1) | reference | 520 (61.0) | reference |
| Anemia: <11g/dL | 82 (29.9) | 1.91 (0.83-4.38) | 333 (39.0) | 2.04 (0.93-4.48) |
| **Referral** | 241 (88.0) | 6.41 (0.82-50.11) | 769 (90.2) | 1.44 (0.46-4.48) |
| **Prior antibiotic use** | 90 (32.9) | 2.13 (0.73-6.22) | 333 (39.0) | 1.50 (0.58-3.86) |
| **Prior antimalarial use** | 4 (1.5) | 2.48 (0.23-26.81) | 29 (3.4) | 1.78 (0.48-6.67) |
| **Respiratory distress** | 66 (24.1) | 1.20 (0.49-2.92) | 162 (19.0) | 1.62 (0.72-3.62) |
| **Maternal and Social Characteristics** |  |  |  |  |
| **Time to reach hospital\*** |  |  |  |  |
| < 30m | 112 (40.9) | reference | 341 (40.0) | reference |
| 30 min - 1h | 88 (32.1) | 1.49 (0.55-4.09) | 350 (41.0) | 1.36 (0.48-3.90) |
| >1h | 74 (27.0) | 0.90 (0.31-2.63) | 162 (19.0) | 3.54 (1.26-9.93) |
| **Water source\*** |  |  |  |  |
| Municipal water/tap | 182 (66.4) | reference | 533 (62.5) | reference |
| Other sources | 92 (33.6) | 0.84 (0.36-1.96) | 320 (37.5) | 2.01 (0.94-4.32) |
| **Boil/disinfect/filter water\*** | 94 (34.3) | 0.21 (0.06-0.72) | 347 (40.7) | 0.74 (0.34-1.62) |
| **Maternal education<sup>2</sup>** |  |  |  |  |
| No school or ≤P3 | 33 (12.0) | reference | 118 (13.8) | reference |
| P4 to P6 | 114 (41.6) | 0.47 (0.17-1.31) | 343 (40.2) | 0.41 (0.15-1.09) |
| S1 to S6 | 103 (37.6) | 0.10 (0.03-0.40) | 325 (38.1) | 0.38 (0.14-1.02) |
| > S6 | 23 (8.4) | 0.15 (0.03-0.83) | 65 (7.6) | 0.09 (0.01-0.76) |
| **Discharge Characteristics** |  |  |  |  |
| **Discharge status** |  |  |  |  |
| Routine discharge | 264 (96.4) | reference | 833 (97.7) | reference |
| Referred to higher level of care | 5 (1.8) | 1.38 (0.14-13.49) | 16 (1.9) | 4.13 (1.05-16.27) |
| Unplanned discharge | 5 (1.8) | 2.40 (0.24-24.39) | 4 (0.5) | - |
| **Length of stay** | 6 (3-9) | 1.04 (1.01-1.08) | 4 (2-7) | 1.08 (1.05-1.12) |
| **Variables collected only for 0-6-month** |  |  |  |  |
| **Fontanelle\*** | 5 (1.8) | 1.73 (0.17-17.19) | - | - |
| **Neonatal jaundice\*** | 15 (5.5) | 2.42 (0.63-9.23) | - | - |
| **Sucking well when breastfeeding\*** | 117 (42.7) | 0.32 (0.13-0.81) | - | - |
| **Duration of present illness\*** |  |  |  |  |
| < 48h | 134 (48.9) | reference | - | - |
| 48h-7d | 104 (38.0) | 2.13 (0.78-5.84) | - | - |
| >7d | 36 (13.1) | 3.57 (1.05-12.18) | - | - |

\* variables used previously in Smart Discharge prediction models; <sup>1</sup> small number represents cutoff for under 6 months; <sup>2</sup> 3 participants reported unknown level of education Abbreviations: OR = odds ratio; IQR = interquartile range; BCS = Blantyre Coma scale; HIV = human immunodeficiency virus; SpO<sub>2</sub> = oxygen saturation

### Post-discharge mortality

The overall rate of post-discharge mortality among those discharged alive was 5.2% (n=58), with a higher cumulative mortality hazard among younger children (**Figure 2**). Post-discharge deaths occurred at a median of 38 days (IQR 16-97.5) and 33 days (IQR 12-76) in the 0–6-and 6–60 months groups, respectively, with most deaths occurring in the hospital (57.1% [n=16] and 70.0% [n=21], respectively) (**Table 2**). For the 0–6 months group, a waz below −3 (aOR 3.31, 95% CI 1.28-8.04) was associated with increased risk of mortality, while higher maternal education (aOR 0.15, 95% CI 0.03-0.85) and use of clean drinking water (aOR 0.21, 95% CI 0.06-0.72) were protective (**Table 1**). In the 6–60 months group, a waz below −3 was associated with significantly increased risk of mortality (aOR 6.52, 95% CI 2.63-16.16), along with an abnormal coma score (aOR 3.28, 95% CI 1.47-7.34), travel time over 1 hour to a healthcare facility (aOR 3.54, 95% CI 1.26-9.93), and the need for referral to higher care (aOR 4.13, 95% CI 1.05-16.27). Higher maternal education was also protective in the 6–60 months group, reducing mortality risk (aOR 0.09, 95% CI 0.01-0.76) (**Table 1**).

**Figure 2:**
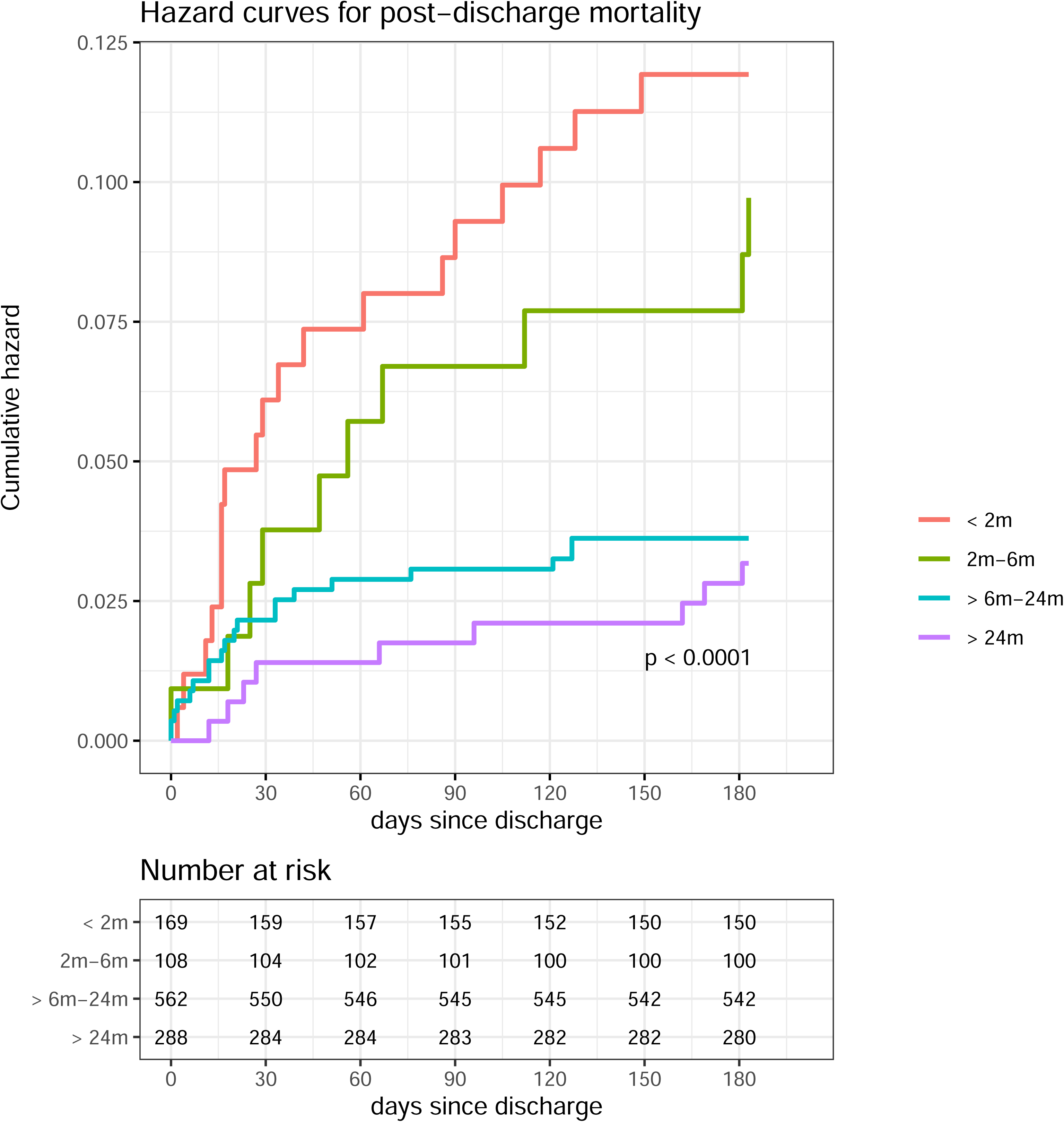
Hazard curves for post-discharge mortality, stratified by age.

**Table 2:**
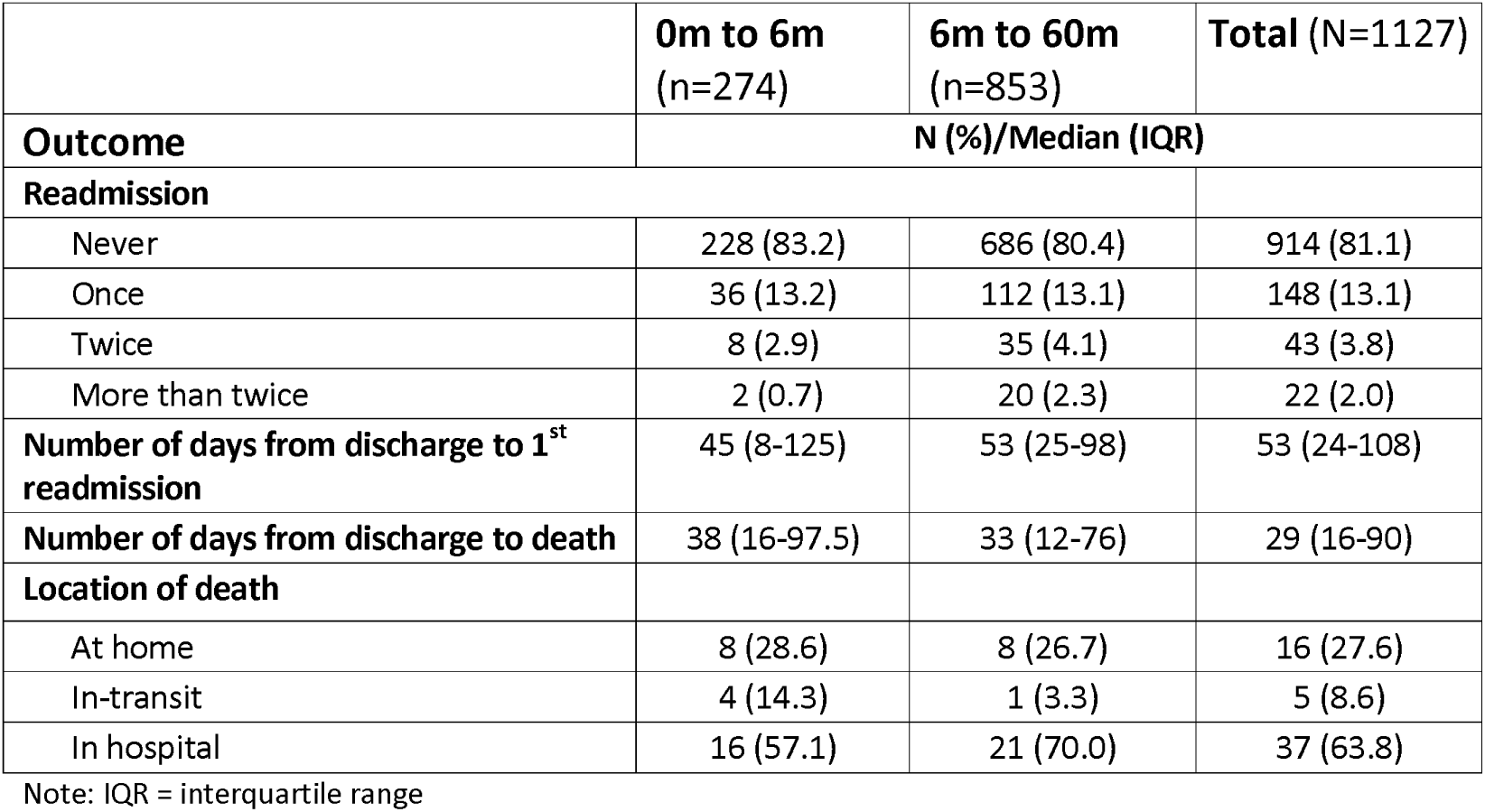
Secondary endpoint and characteristics of post-discharge deaths.

|  | 0m to 6m<br>(n=274) | 6m to 60m<br>(n=853) | Total (N=1127) |
| --- | --- | --- | --- |
| Outcome | N (%) / Median (IQR) |  |  |
| Readmission |  |  |  |
| Never | 228 (83.2) | 686 (80.4) | 914 (81.1) |
| Once | 36 (13.2) | 112 (13.1) | 148 (13.1) |
| Twice | 8 (2.9) | 35 (4.1) | 43 (3.8) |
| More than twice | 2 (0.7) | 20 (2.3) | 22 (2.0) |
| Number of days from discharge to 1<sup>st</sup> readmission | 45 (8-125) | 53 (25-98) | 53 (24-108) |
| Number of days from discharge to death | 38 (16-97.5) | 33 (12-76) | 29 (16-90) |
| Location of death |  |  |  |
| At home | 8 (28.6) | 8 (26.7) | 16 (27.6) |
| In-transit | 4 (14.3) | 1 (3.3) | 5 (8.6) |
| In hospital | 16 (57.1) | 21 (70.0) | 37 (63.8) |

Note: IQR = interquartile range

### Post-discharge readmission

The overall rate of post-discharge readmission was 18.9% (n=213), with 5.8% (n=65) of children experiencing multiple readmissions (**Table 2**). The median time to first readmission was 45 days (IQR 8-125) and 53 days (IQR 25-98) for the 0–6 and 6–60 month groups, respectively. Unlike post-discharge mortality, readmission was not significantly affected by age, although there was a trend suggesting that those younger than 2 months may have a lower risk of readmission (**Figure 3**).

**Figure 3:**
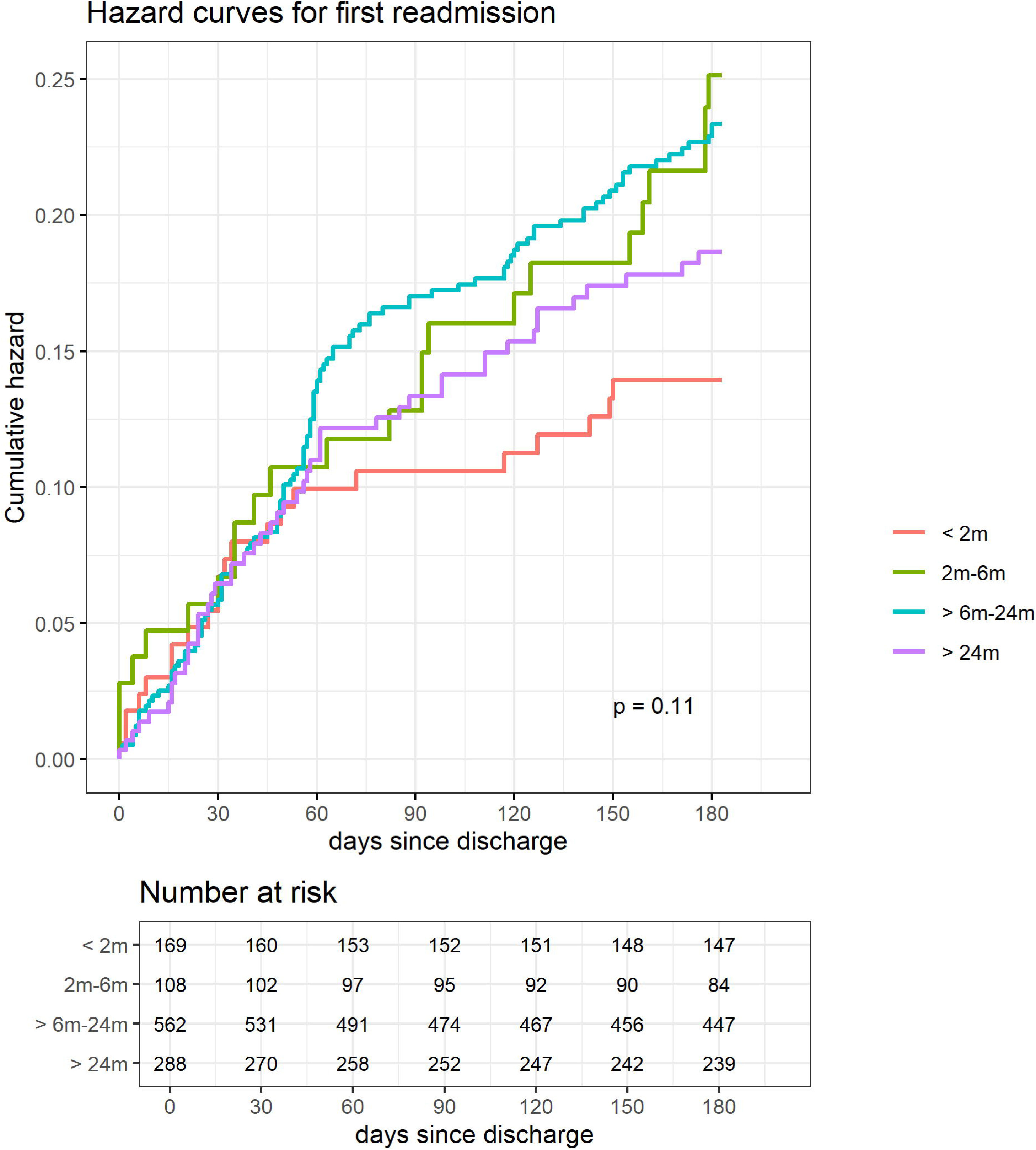
Hazard curves for first readmission, stratified by age.

## Discussion

In our prospective observational study of children under five admitted with suspected or confirmed infections in Rwanda, nearly 1 in 20 children died after discharge, similar to the rate of mortality during the index hospital admission. We found key clinical and socio-behavioral factors are associated with higher odds of post-discharge mortality, including, for both age groups, severe malnutrition (WAZ < −3); and, for children aged 6–60 months, malnutrition, abnormal coma scores, long travel times to healthcare, and the need for referral to higher-level of care. Infants aged < 2 months had the highest risk of death.

These results as well as the risk factors identified are largely in line with previous studies conducted in Uganda and elsewhere in East Africa [3,22,23]. The negative health effects of malnutrition are well known and include reduced immune competence [24,25] and deficiencies in macro- and micronutrients [26,27], leading to a cycle of recurrent infections and deteriorating nutritional status, which further increases the risk of post-discharge mortality [28]. We also found higher post-discharge mortality rates in urban Kigali (9%) than in rural Ruhengeri (3%), in contrast to the typical rural-urban health disparity in many LMICs, where patients in rural areas typically fare worse due to limited healthcare access and resources [29].

We hypothesize that elevated mortality rates in Kigali are likely due to CHUK’s role as a tertiary care hospital admitting the most critically ill pediatric patients nationwide, including referrals of severe and complex cases from Ruhengeri, which does not have ICU capacity.

Rwanda has already implemented programs to improve child health and is one of the few countries in sub-Saharan Africa to have achieved the Millennium Development Goal (MDG) related to under-5 mortality [30]. With several key health systems measures already well established in Rwanda, the use of simple models to identify the “at risk” child could be leveraged towards an effective solutions to address the high rates of post-discharge mortality [31]. These include programs such as the well-established Community Health Worker (CHW) program for follow-up care [32], “Mutuelle de Santé,” a community-based health insurance program, which lowers financial barriers to potentially improve access to post-discharge services [33], a unified Health Management Information System (HMIS) to facilitate patient tracking and widespread integration of risk-based prediction models [34], and the Mentoring and Enhanced Supervision at Health Centers (MESH) for healthcare providers that could be used for training and implementation of system-wide post-discharge care and education packages[35]. These systems argue that more immediate implementation of discharge education and community-based interventions may have substantial effects [36–38] on outcomes. Other public health campaigns and community-based programs, increased access to quality education, nutritional supplementation, expanded access to healthcare, and imprpoved socioeconomic conditions, would also help reduce both post-discharge and overall child mortality in Rwanda [39].

### Limitations

The limitations of this study include a small sample from only two hospitals, potentially limiting its generalizability to other regions in Rwanda or similar settings, and a six-month observation period possibly missing longer-term effects. The primary data collection method and interviews may have introduced recall bias and inaccuracies. Despite this, the study provided detailed information on the severity of the children’s conditions and comorbidities. The study did not fully explore all socioeconomic and environmental factors, healthcare quality, disease severity, concurrent illnesses, genetic influences, and healthcare-seeking behaviors affecting post-discharge mortality. Future research should address these limitations by using a larger, more diverse sample, extending the follow-up period, and conducting a comprehensive analysis of the relevant factors.

### Conclusions

This study identified a significant burden of post-discharge mortality among pediatric patients in Rwanda, particularly affecting infants and those who are socially vulnerable, including those with malnutrition. These findings underscore the need for targeted interventions to address risk factors, improve healthcare access, and enhance care during the post-discharge period. This study also calls for more longitudinal research to identify additional factors influencing post-discharge mortality and the development of interventions and implementation strategies. A comprehensive approach that combines improvements in the health system with broader socioeconomic initiatives is needed to reduce post-discharge mortality and improve pediatric health outcomes.

## Data Availability

All data produced are available online through the Smart Discharges Dataverse in deidentified format.

https://borealisdata.ca/dataset.xhtml?persistentId=doi:10.5683/SP3/60DTRF

## Acknowledgements

We would like to acknowledge all past and present members of the Smart Discharges Research program for their efforts in data collection, administration, logistics support, and all study activities, including but not limited to: Godefroid Rucinga, Esperance Umulisa, Didas Mugambinumwe, Jeanne d’Arc Mazimpaka, Claudine Uwingabiye, Theogene Bizimungu, Juliette Unyuzumutima, Peter Lewis, and Martina Knappett.

## Supplementary Files

**Table S1:** Supplementary Table 1, Overall demographics and cohort characteristics.

|  | **N = 1127** |  |
| --- | --- | --- |
| **Variables** | **N (%) / Median (IQR)** | **aOR\* (95% CI)** |
| **Site, n (%)** |  |  |
| Kigali | 361 (32.3) | reference |
| Ruhengeri | 766 (68.0) | 0.24 (0.14-0.43) |
| **Male (ref=female)** | 676 (60.0) | 0.81 (0.47-1.39) |
| **Age, months** | 13.5 (6.1-24.7) | 0.96 (0.94-0.99) |
| **MUAC (mm)<sup>1</sup>** |  |  |
| <110 / <115 | 109 (9.7) | 2.69 (1.19-6.08) |
| 110-120 / 115-125 | 112 (9.9) | 3.01 (1.47-6.17) |
| >120 / >125 | 906 (80.4) | reference |
| **Weight for age z-score** |  |  |
| <-3 | 103 (9.1) | 4.63 (2.42-8.85) |
| -3 to -2 | 116 (10.3) | 2.62 (1.22-5.65) |
| >-2 | 908 (80.6) | reference |
| **SpO<sub>2</sub>, %** | 95 (88-97) | 0.98 (0.95-1.02) |
| **Heart rate** | 142 (128-157) | 1.00 (0.99-1.02) |
| **Respiratory rate** | 41 (35-49) | 1.01 (0.99-1.03) |
| **Temperature** |  |  |
| < 36.5 | 208 (18.5) | 0.75 (0.34-1.66) |
| 36.5-37.5 | 419 (37.2) | reference |
| >37.5 | 500 (44.4) | 0.67 (0.37-1.22) |
| **Abnormal BCS** | 194 (17.2) | 1.93 (1.06-3.52) |
| **HIV positive** | 3 (0.3) | 4.04 (0.35-46.89) |
| **Positive malaria test** | 17 (1.5) | - |
| **Haemoglobin, g/dl** |  |  |
| No anemia: ≥ 11 | 712 (63.2) | reference |
| Anemic: < 11 | 415 (36.8) | 1.74 (1.00-3.00) |
| **Respiratory distress** | 228 (20.2) | 1.37 (0.76-2.48) |
| **Referral** | 1010 (89.6) | 2.37 (0.90-6.24) |
| **Prior antibiotic use** | 423 (37.5) | 1.58 (0.80-3.11) |
| **Prior antimalarial use** | 33 (2.9) | 1.98 (0.63-6.23) |
| **Maternal and Social Characteristics** |  |  |
| **Time to reach hospital** |  |  |
| <30 min | 453 (40.2) | reference |
| 30 min - 1h | 438 (38.9) | 1.28 (0.63-2.60) |
| >1h | 236 (20.9) | 1.88 (0.91-3.88) |
| **Maternal education<sup>2</sup>** |  |  |
| No school or ≤P3 | 151 (13.4) | reference |
| P4 to P6 | 457 (40.6) | 0.46 (0.23-0.91) |
| S1 to S6 | 428 (38.0) | 0.24 (0.11-0.52) |
| > S6 | 88 (7.8) | 0.13 (0.04-0.49) |
| **Water source** |  |  |
| Municipal water/tap | 715 (63.4) | reference |
| Other sources | 412 (36.6) | 1.33 (0.76-2.30) |
| **Boil/disinfect/filter water** | 441 (39.1) | 0.46 (0.24-0.88) |
| **Discharge Characteristics** |  |  |

| **Discharge status** |  |  |
| --- | --- | --- |
| Routine discharge | 1097 (97.3) | reference |
| Referred to higher level of care | 21 (1.9) | 2.77 (0.85-9.01) |
| Unplanned discharge | 9 (0.8) | 1.57 (0.18-13.50) |
| **Length of stay** | 4 (3-8) | 1.06 (1.04-1.09) |

Note: \*adjusted for age, sex, and site. <sup>1</sup>small number represents cutoff for under 6 months; <sup>2</sup>3 participants reported unknown level of education
Abbreviations: OR = odds ratio; IQR = interquartile range; BCS = Blantyre Coma scale; HIV = human immunodeficiency virus; SpO<sub>2</sub> = oxygen saturation

